# Intraoperative ultrasound localization microscopy of human brain tumors and arteriovenous malformations

**DOI:** 10.1101/2025.09.19.25335978

**Authors:** Luxi Wei, Luuk Verhoef, Paul Xing, Emma C. Gommers, Sadaf Soloukey, Marion Smits, Wouter B.L. van den Bossche, Victor Volovici, Clemens M.F. Dirven, Joost W. Schouten, Eelke M. Bos, Anne Kleijn, Peter P. de Smalen, Arnaud J.P.E. Vincent, Pieter Kruizinga

## Abstract

The microvasculature and hemodynamics of human brain tumors and other lesions have remained largely unexplored *in vivo* due to the limited resolution of conventional imaging techniques, and may present new opportunities in biomarker identification and neurosurgeries. Ultrasound Localization Microscopy (ULM), which tracks freely circulating intravascular microbubbles used as contrast agents, overcomes these limitations and has been used to visualize vascular structures and flow. In this study, we performed intraoperative ULM during brain surgeries involving resection of brain tumors (N = 3 meningiomas, N = 3 brain metastasis, N = 3 high grade gliomas) and an arteriovenous malformation (AVM) (N = 1). ULM provided microvascular images of the human brain, resolving vessels down to 35 μm, revealing clear differences in structure and dynamics between tumor and surrounding healthy tissue. By following individual microbubbles, vessel connectivity was probed and used to identify feeding, draining, and non-tumoral vessels. In one case, a 3D ULM map of an AVM was generated with 226 μm resolution, allowing us to resolve its complex internal structure, including feeding and draining vessels. Intraoperative ULM enables visualization of human brain vasculature and hemodynamics at unprecedented resolutions, and may directly aid tumor and AVM resections by providing flow paths and speeds through compact niduses.

**One Sentence Summary:** Ultrasound localization microscopy during human brain surgery revealed vascular structure and dynamics of brain lesions at sub-millimeter resolution.

## INTRODUCTION

Our understanding of the microvascular structure and dynamics of the human brain remains fundamentally limited; it’s primarily derived from post-mortem microscopy techniques and extrapolations from small animal models. While advanced imaging approaches—such as light sheet microscopy in cleared rodent brains (1) and sophisticated X-ray methods applied to post-mortem human tissue (2)—have yielded valuable insights into vascular architecture, significant gaps persist in our knowledge of the human brain *in vivo*, particularly regarding the underlying hemodynamics that govern blood flow at millimeter and sub-millimeter scales. The literature reveals a striking absence of data on actual flow velocities within deep brain nuclei, cortical arterioles and venules, and the microvascular territories critical to neural function (3). This deficit extends to pathological conditions. Pathological angiogenesis in high-grade glioma, meningioma, and brain metastasis produces distinct microvascular signatures marked by altered vessel tortuosity, abnormal branching, and disrupted flow dynamics (4), while arteriovenous malformations (AVMs) create 3-dimensional altered-flow, high-risk circuits that may contain compact niduses prone to rupture (5)—yet the precise microscale hemodynamics of these lesions remain poorly characterized.

Access to high-resolution microvascular structure and hemodynamic information could fundamentally transform our understanding of human cerebral dynamics in health and disease, and in turn potentially trigger new strategies for disease treatment and follow-up. Previous studies have suggested that assessment of tumor vascularity has the potential to improve prognostic precision, help discover potential molecular therapies, and can be used to assess the results of anti-tumor therapies (6–9). Blood flow information at the microvascular level can potentially open new avenues for the discovery of vascular biomarkers to improve tumor characterization (10–12). In the clinical context of neurosurgery, hemodynamic information can be used to better predict the amount of blood loss during tumor resection and vascular surgeries, potentially informing both intraoperative decision-making and post-surgical treatment approaches (13–15).

Current clinical imaging modalities cannot bridge this critical gap in resolution. Digital subtraction angiography (DSA), the gold standard for vascular visualization, requires ionizing radiation and cannot resolve vessels much smaller than 1mm (16,17). The low frame rate (10-30 Hz) also limits its use to in and outflow of organs or vascular abnormalities, incapable of capturing blood flow speeds (16). Indocyanine green (ICG) videoangiography provides real-time fluorescent visualization but suffers from limited depth penetration and similar resolution constraints (18,19). Magnetic resonance imaging (MRI) avoids harmful radiation and depth limitations but lacks the spatial (∼1 mm) necessary to visualize microvasculature and its flow patterns (20,21). The most advanced high-field MRI scanners offer excellent pial vessel visualization but cannot resolve arterioles and venules where critical pathological processes occur (22). Intraoperatively, MRI-machines come with an additional burden of severe disruption of the surgical workflow. These limitations leave critical vascular territories invisible to surgeons.

Ultrasound localization microscopy (ULM) represents a transformative approach capable of exploring this previously inaccessible territory (23,24). ULM overcomes the fundamental diffraction limit of conventional ultrasound through tracking individual microbubble contrast agents flowing through microvasculature. This achieves super-resolution vascular mapping with spatial resolution approaching 10-20 micrometers—an order of magnitude improvement over conventional ultrasound’s 200-500 micrometer resolution (25). This breakthrough parallels the revolutionary optical super-resolution microscopy techniques recognized by the 2014 Nobel Prize in Chemistry (26,27).

Translation from preclinical research to clinical application has faced challenges including patient motion, skull bone interfaces, and demanding real-time processing requirements (28–31). However, a unique opportunity emerges during neurosurgical interventions. Craniotomy provides ultrasound with direct brain tissue access, eliminating skull-related attenuation and artifacts. The ultrasound probe can be fixed in place, enabling stabilized, minute-long recordings needed to reach high resolutions. Previously, B-mode and ultrafast Doppler ultrasound have been performed under this intraoperative setting and achieved visualization of hemodynamic features including flow velocities, directional patterns, and functional signals in living human brain tissue (25,32), but not to the same spatial resolution achievable with ULM because of the ultrasound diffraction limit (33).

In this study, we report the first systematic clinical application of intraoperative ULM during neurosurgical procedures in nine patients with brain tumors (meningioma, metastasis, and high-grade glioma). We achieved visualization of microvasculature down to 35 μm—previously attainable only via post-mortem histology—and provided the first quantitative estimates of blood flow velocities in cortical and striatal territories. We demonstrated significant differences in microvascular speed and tortuosity between healthy and diseased tissue, highlighting the potential to identify additional biomarkers at high resolution. These results, obtained using a conventional large-aperture linear probe, enable detailed study of human brain structure and dynamics, while the use of standard linear probe facilitates reproducibility across surgical centers. Additionally, we present the first 3D ULM reconstruction in a human AVM case— achieved using a smaller-aperture 2D experimental matrix probe—resulted in reduced resolution (226 μm) yet sufficient to reveal flow through the AVM nidus with detail beyond conventional imaging.

## RESULTS

### ULM can be performed in an intraoperative setting

Between April 2024 and March 2025, 10 patients (5 females, 5 males; mean age of 64 (46–79) years) were enrolled in this study (see Table S1). Diagnoses included metastasis (n = 3, patient #1-3), high-grade glioma (n = 3, patient #4-6), meningioma (n = 3, patient #7-9), and AVM (n = 1). In all cases, the lesions were clearly visualized using conventional gray-scale ultrasound (B-mode) and intraoperative contrast-enhanced ultrasound. No surgical complications occurred.

Figure 1 provides an overview of the intraoperative ULM methodology and its capabilities. Following craniotomy and prior to durotomy, the ultrasound probe was positioned and fixed above the tumor area, coupled to the dura with saline (see Methods: Study procedure). Fig. 1A shows a schematic of the setup. Intravenous injection of the ultrasound contrast agent was synchronized with the start of ultrafast data acquisition, and image acquisition and beamforming were performed in real time using custom software. The beamformed images were then processed using a modified version of the PALA toolbox (34), and key steps are outlined in Fig. 1B. In each frame, individual microbubbles were first localized via Gaussian fitting, then tracked spatiotemporally across acquisitions to reconstruct flow trajectories. Cumulative tracking over time yielded super-resolved vascular maps that reveal sub-diffraction-scale vessel architecture and directional flow dynamics (24). Fig. 1C illustrates the diverse vascular insights enabled by ULM beyond conventional ultrasound imaging. In addition to standard B-mode and power Doppler images (PDI), ULM resolves vessel architecture, flow velocity, and direction at the microvascular scale. By following the movement of individual microbubbles, the connectivity of vascular networks can also be reconstructed. Additionally, the temporal resolution of minute-long acquisitions allows for analysis of flow dynamics across cardiac cycles. The following sections demonstrate these capabilities in human brain tumors and AVM.

**Fig. 1.**
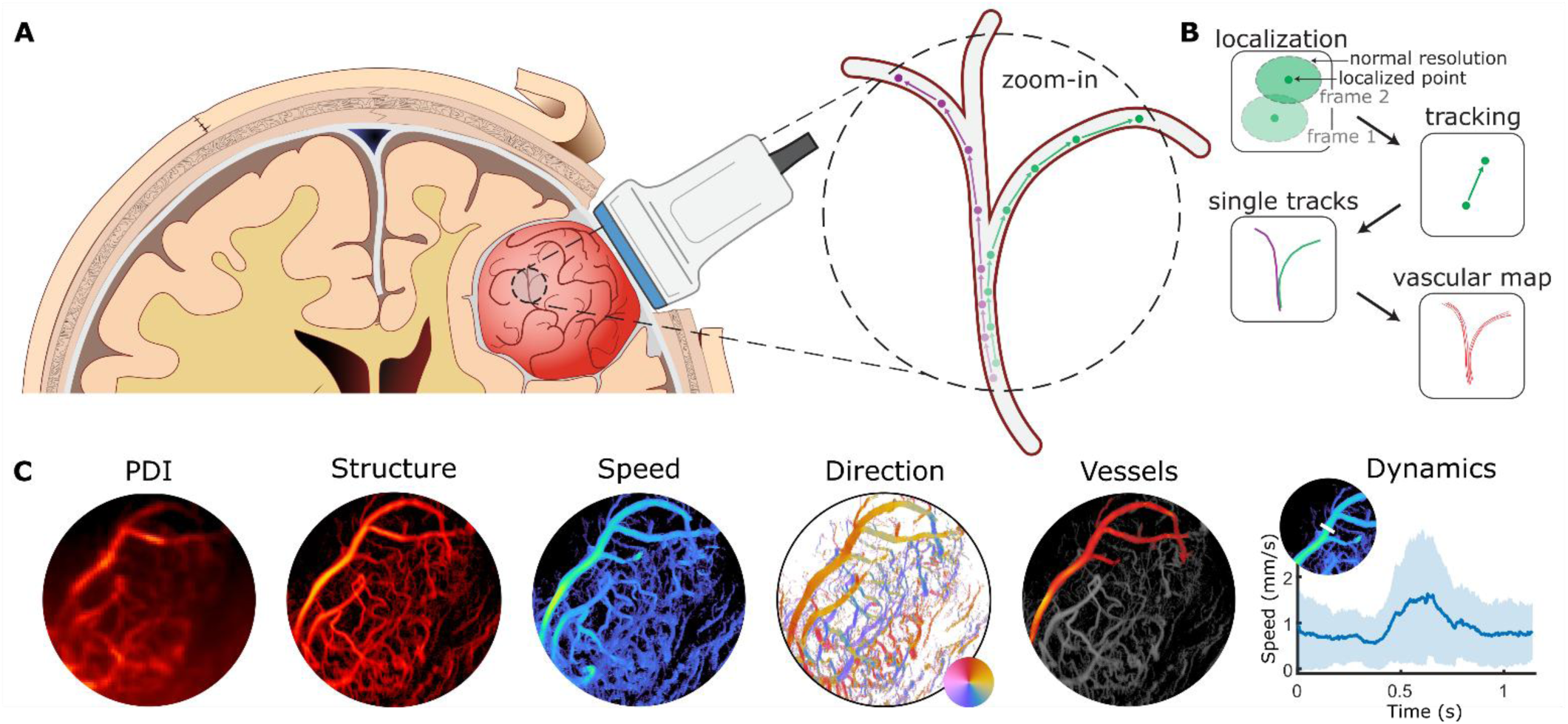
Ultrafast contrast-enhanced ultrasound imaging during brain surgery enables ULM capabilities. **(A)** Schematic of the experimental setup. During human brain tumor resections, the ultrasound probe was placed in contact with the dura immediately following craniotomy. Ultrafast ultrasound imaging was used to capture intravenously injected microbubbles as they traversed the vascular network. **(B)** ULM post-processing pipeline. This includes microbubble localization and tracking to generate individual trajectories, which are then accumulated and rasterized to create high-resolution vascular maps. **(C)** In addition to conventional B-mode and power Doppler imaging (PDI), ULM processing enables sub-millimeter resolution visualization of vascular structures, along with blood flow speed and direction. Individual vessels can be segmented based on track connectivity, and flow dynamics can be assessed across the cardiac cycle.

### Intraoperative ULM reveals microvascular structures and dynamics of human brain tumors

We analyzed microvascular features—including vessel patterns, tortuosity, flow speed, and connectivity—in representative cases of metastasis, high-grade glioma, and meningioma to demonstrate the capabilities of intraoperative ULM for characterizing tumor-specific vascular parameters and its potential to guide surgical dissection. Fig. 2 presents a detailed analysis of a urothelial cell carcinoma metastasis (patient #1), located in the left parietal lobe (see Fig. S1E and F). Fig. 2A shows the ULM density-based map, where pixel intensity reflects the number of microbubbles passing through each location. Notably, the directionality and tortuosity of microvasculature within the tumor qualitatively differ from the surrounding tissue, delineating a clear tumor boundary (green arrowheads). Beneath the lesion, healthy vessels appear deflected away from the tumor mass (turquoise arrowhead), likely due to mechanical displacement caused by tumor expansion. The resolution of this image was calculated to be 37 µm using the Fourier

**Fig. 2.**
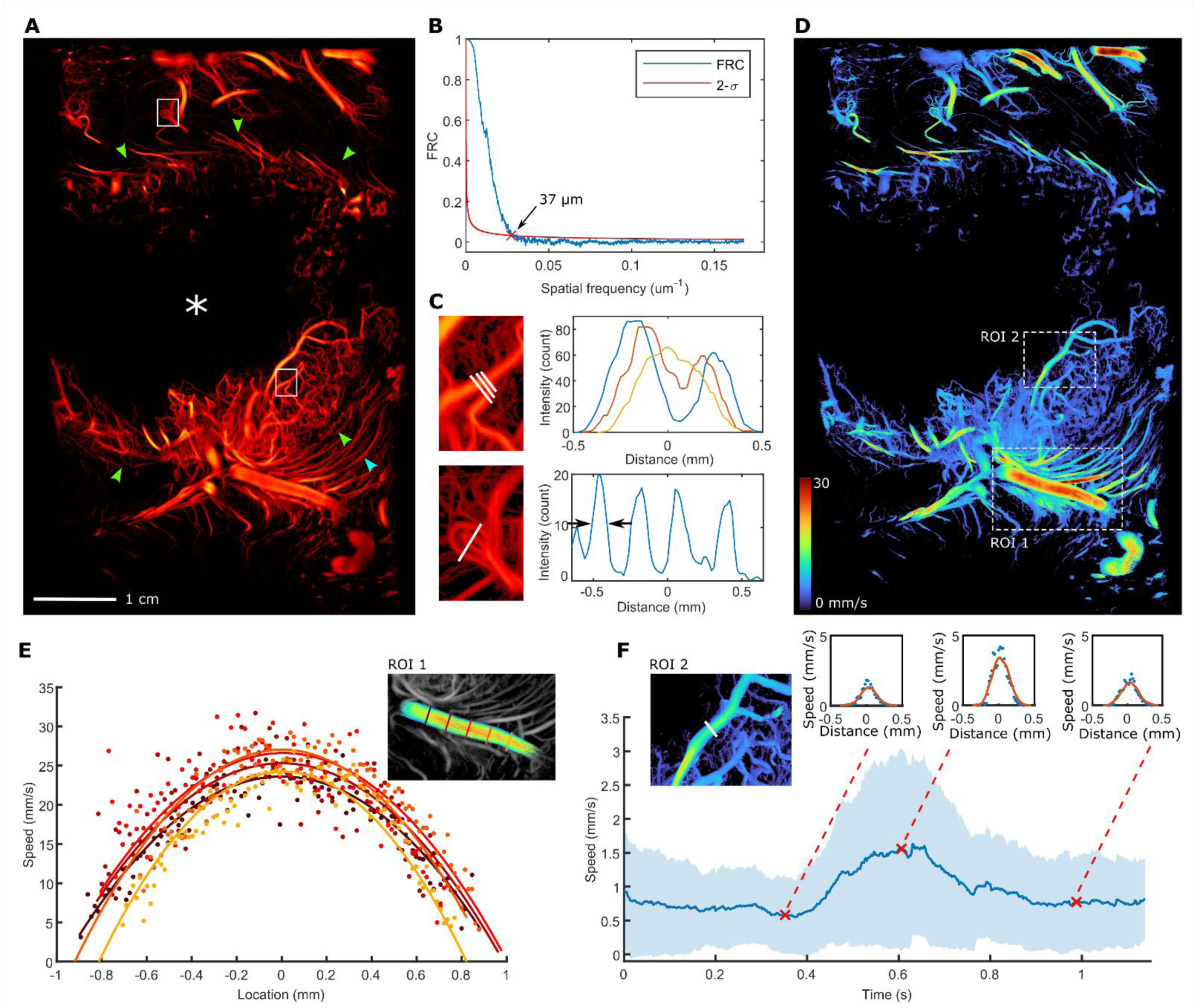
ULM reveals microvasculature structure and dynamics. **(A)** ULM density-based render of a brain metastasis (patient #1). An avascular necrotic core (white asterisk), the tumor border (green arrowheads), and deformed surrounding vasculature (turquoise arrowhead) are visible. **(B)** The spatial resolution of (A) was estimated to be 37 um using the Fourier Ring Correlation (FRC) method. **(C)** Zoomed-in views and cross-sectional intensity profiles of two regions of interest (white boxes in A) demonstrate the sub-millimeter resolution achievable with this technique. The full-width-half-maximum of the four vessel cross-sections (black arrows) are calculated to be 0.11 ± 0.01 mm. **(D)** Blood flow speeds were calculated and color-coded across the field of view. Two boxed regions are selected for further analysis in panels (E) and (F). **(E)** A large vessel at the base of the tumor was isolated, and the flow speeds across its diameter were calculated at five positions (see inset) during diastole. Parabolic flow profiles were observed at all locations (least-squares fit), consistent with laminar flow. **(F)** The mean and standard deviation of blood flow speeds along the white line in (D) were computed across a full cardiac cycle. Flow profiles at each time point of the cycle can also be extracted, as illustrated by the insets.

Ring Correlation (FRC) method (35) (Fig. 2B), allowing us to observe microvascular features that were never before resolved *in vivo* previously. Fig. 2C highlights two such examples from the white-boxed areas in Fig. 2A: a sub-millimeter vessel bifurcation within the tumor (top), and a looped vessel structure outside of the tumor (bottom). The latter may be the result of tumor-induced remodeling (12,36,37).

In addition to visualizing sub-millimeter vessel structures, ULM can be used to measure blood flow speed. Fig. 2D presents the average flow speed map for this tumor. Larger vessels exhibited higher flow speeds, both within and outside of the tumor. Zooming into the two boxed regions in Fig. 2D, Fig. 2E shows velocity profiles measured at five equally spaced locations across a large draining vessel during diastole (see inset). Parabolic profiles were observed during diastole, consistent with laminar flow, with a maximum flow speed of 27 mm/s (Fig. 2E). Blood flow speed during systole within this large vessel was too high (max detectable was 51 mm/s) to obtain accurate measurements. Finally, using the method described in Ghigo *et al* (38), the entire ULM recording was synchronized to one cardiac cycle, allowing pulsatile flow dynamics to be analyzed for an example vessel (Fig. 2F). Along the white line shown in the Fig. 2F inset, the mean flow speed varied across the cardiac cycle according to the expected pattern. At three example timepoints during the cardiac cycle, the flow profiles were plotted (Fig. 2F insets), demonstrating the ability to capture pulsatile flow patterns of small vessels.

A second tumor type, a high-grade glioma (patient #4) located in the left occipital lobe (Fig. S1 C, D), is illustrated in Fig. 3. Fig. 3A shows the ULM density-based map, where the microvasculature of the tumor and the surroundings could be visualized. A large hypo-vascular necrotic core (white asterisk) was found at the center of the tumor, in agreement with pre-operative MRI results (Fig. S1 C, D). The tumor border could be identified by a change in B-mode intensity and vascular structure (Fig. 3B, yellow dashed line). Interestingly, vessel-like structures were visible on the B-mode image within the necrotic core, although some of these showed no blood flow in the ULM map, indicating possible non-perfused vessels or vessel-like structures. Structural vessel modifications were evident at the tumor border, where they became denser and more tortuous (Fig. 3A, C), consistent with known characteristics of invasive tumors such as high-grade glioma (9,36,39). To assess these changes quantitatively, we defined two concentric ring-shaped regions-of-interest (ROIs) located directly inside and outside the segmented tumor border. Vessel tortuosity was evaluated using the distance metric (DM) (40), overlaid on the ULM density-based map in Fig. 3C.

**Fig. 3.**
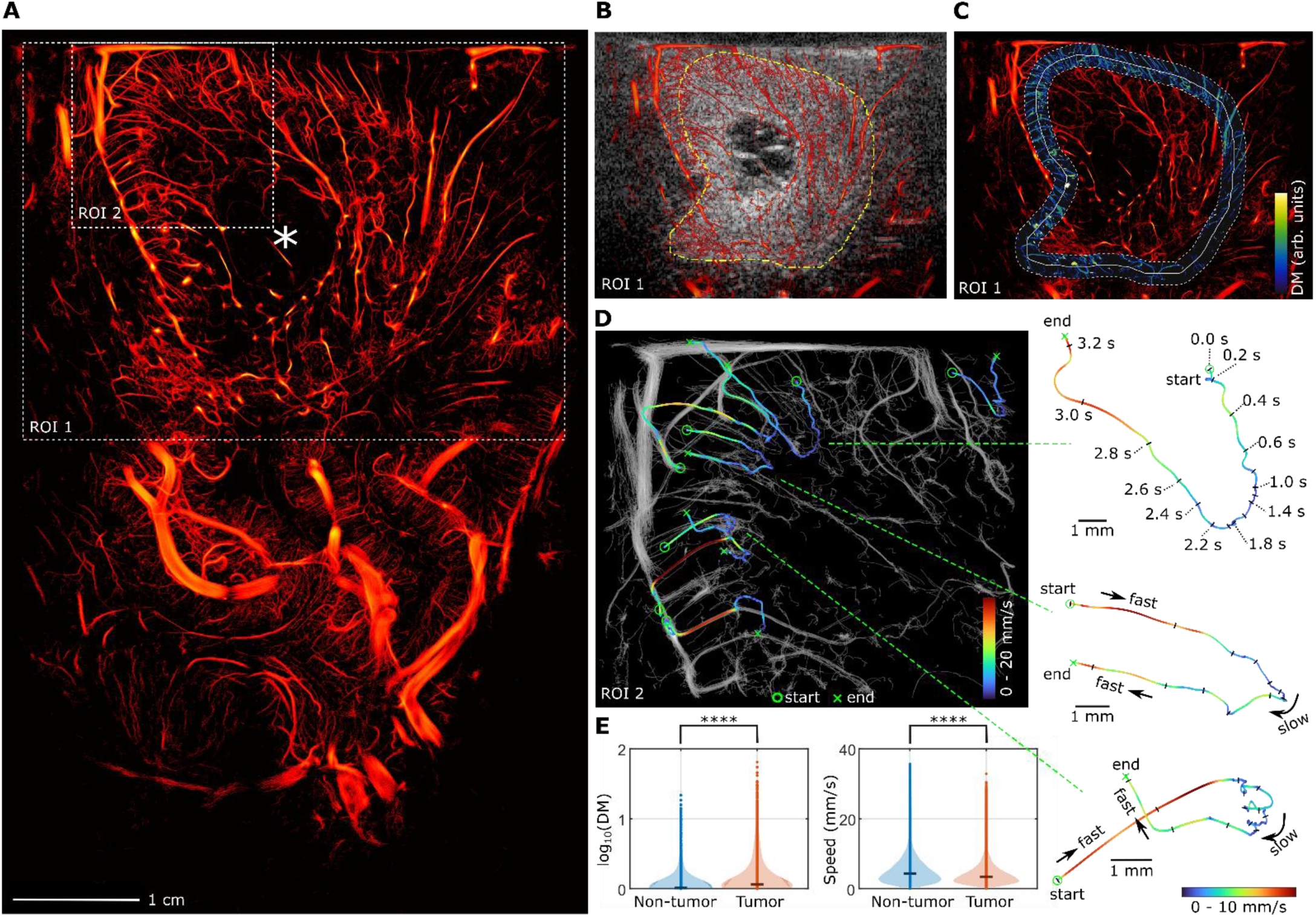
Tumor characteristics can be identified using single microbubble tracks. **(A)** ULM density-based image of a high-grade glioma (patient #4). The non-vascular nechrotic core is labeled (white asterisk). **(B)** ULM density map overlaid on the B-mode image of ROI1 in (A). The border of the tumor (yellow dashed line, drawn using the B-mode image) can be distinguished both as an intenisty change in the B-mode image and a microvascular structure change in the ULM density image. **(C)** Vessel tortuosity, quantified using the distance metric (DM), is mapped near the tumor border. The white solid line indicates the tumor boundary, while the white dashed lines represent regions immediately inside and outside the tumor. These two regions are used to distinguish tumor and non-tumor revions in (E). **(D)** Shows a zoomed-in render of ROI2, labeled with example single microbubble tracks. These microbubbles can be seen moving into and draining out of the tumor (circle: start; cross: end). Their flow speed changes along their paths. **(E)** Violinplots showing comparisons of DM values and microbubble speeds between tumor and non-tumor regions adjacent to the tumor border (as defined in panel C). Statistically significant differences were observed (****P < 0.001, n_non-tumor_ =163107, n_tumor_ = 218799, Wilcoxon rank-sum test).

Structural and hemodynamics differences were observed at the tumor border region between the tumor and surrounding tissue. Single microbubbles could be identified and tracked as they entered the tumor, slowed down, and exited (Fig. 3D). Using the two ROIs from Fig. 3C, we calculated relative blood volume (rBV), DM, and microbubble speed to compare vascular characteristics across the tumor border. The rBV values were 36% inside and 24% outside the tumor border. Distributions of DM and microbubble speed within each ROI are shown in Fig. 3E. Vessels inside the tumor border were significantly more tortuous, with median DM values of 1.14 compared to 1.02 outside of the tumor (p < 0.001, Wilcoxon rank-sum test). Conversely, microbubble speeds were significantly higher outside the tumor border, with median values of 4.3 mm/s versus 3.4 mm/s inside the tumor (p < 0.001, Wilcoxon rank-sum test).

A different tumor type with low aggressiveness (grade 1 meningioma), located extra-axially in patient #9, is visualized in Fig. 4. The ULM density-based map (Fig. 4A) reveals a vascular pattern characteristic of meningioma, where vessels originate from and drain into the meninges (41,42). This pattern is even more evident in the flow direction map of Fig. 4B, where most vessels exhibit flow either toward or away from the meningeal surface at the top of the image. To further analyze the feeding and draining vascular structures, we followed and grouped microbubble trajectories that belonged to the same vessels to reconstruct individual vascular trees, beginning at their trunks near the meningeal surface (Fig. 4C and D). Using this approach, connected vessels were isolated and grouped into arterial and venous trees. ROI1 in Fig. 4A, focused on the tumor region, is shown in Fig. 4C with overlaid examples of selected feeding (red) and draining (blue) vascular trees. Fig. 4D are two enlarged examples of such vascular trees. For the feeding vessel, a single vessel was found to form a complex network of microvascular branches, with two instances of vessels reversing direction and flowing back towards the meninges (Fig. 4D, black arrows). For the example draining vessel tree, multiple draining locations at the meninges were observed.

**Fig. 4.**
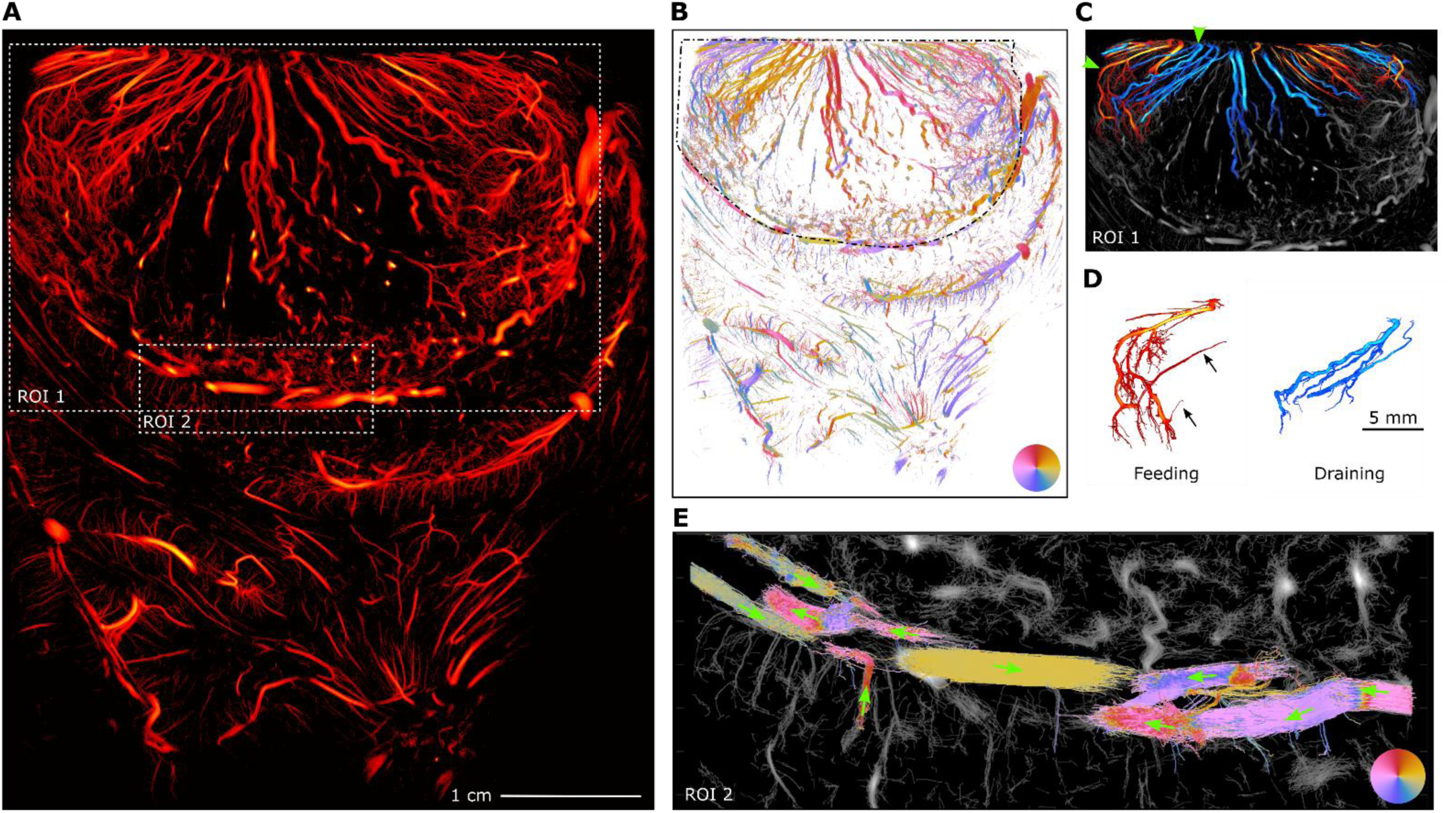
Microbubble tracks can be used to identify connectivity of blood vessels and vascular trees. **(A)** ULM density-based image of a meningioma (patient #9). **(B)** ULM flow direction map. The border of the tumor is labeled with a black dashed line. **(C)** Feeding (red) and draining (blue) vessel trees overlayed on an ULM density map of region-of-interest ROI1 depicted in (A). **(D)** Two example vascular trees (green arrowheads in (C)) are isolated and displayed. **(E)** Flow directions for vessels at the tumor border in ROI2 depicted (A).

Bubble tracking further enabled analysis of vessel connectivity in the peritumoral region. In Fig. 4E, tracks of microbubbles traversing vessels at the base of the tumor were visualized. Trajectory analysis revealed that these vessels did not converge toward or diverge from the tumor within the imaged plane, suggesting they were not functionally integrated into the tumor vasculature. This finding aligned with the surgeon’s intraoperative observation that the tumor was well-separated from the surrounding brain tissue, making it easier to remove with minimal blood loss.

Analyses across the three tumor types demonstrated the capacity of intraoperative ULM to provide high-resolution visualization and quantitative characterization of tumor-associated microvasculature. ULM enables measurement of vessel density, tortuosity, flow dynamics, and vascular remodeling, parameters that may serve as potential biomarkers for tumor identification and grading. ULM also allows for detailed analysis of blood flow velocity and vessel connectivity. This information could potentially inform surgeons about anticipated blood loss during resection, supporting intraoperative decision-making and enabling safer tumor removal.

### *In vivo* characterization of healthy human brain vasculature

While imaging brain tumors, we were able to capture interesting structures and dynamics in the surrounding brain regions at scales that were previously accessible only through post-mortem techniques. Fig. 5A-D shows MRI and ULM results from patient #4, diagnosed with a high-grade glioma, along with a microscopy reference image from (43). In Fig. 5A, the red box ROI labels displaced cortical gray matter underneath the tumor on a T1-weighted MRI. Pial vessels found within the sulcus (Fig. 5B, green arrowheads) and appeared to make a sharp turn at the base of the sulcus (Fig 5C, yellow arrowhead). From the pial vessels, smaller, parallel cortical penetrating arterioles and venules perfused the cortical gray matter. Additionally, deep cortical branches were observed to extend beyond the cortical layer and turn upon entry into the white matter layer (yellow arrowheads). Flow directions in these vessels were captured using direction-encoded ULM imaging (Fig. 5C), with a reference line drawn along the intercortical vessel (yellow dashed line); blood flow directed away from and toward this reference was color-coded in green and purple, respectively. Median blood flow speeds were measured to range from 2.5 mm/s in the cortical penetrating branches and 10.4 mm/s in the larger pial vessels. These structural observations were consistent with that of the reference (Fig. 5D) as well as other existing literature (44–46).

**Fig. 5.**
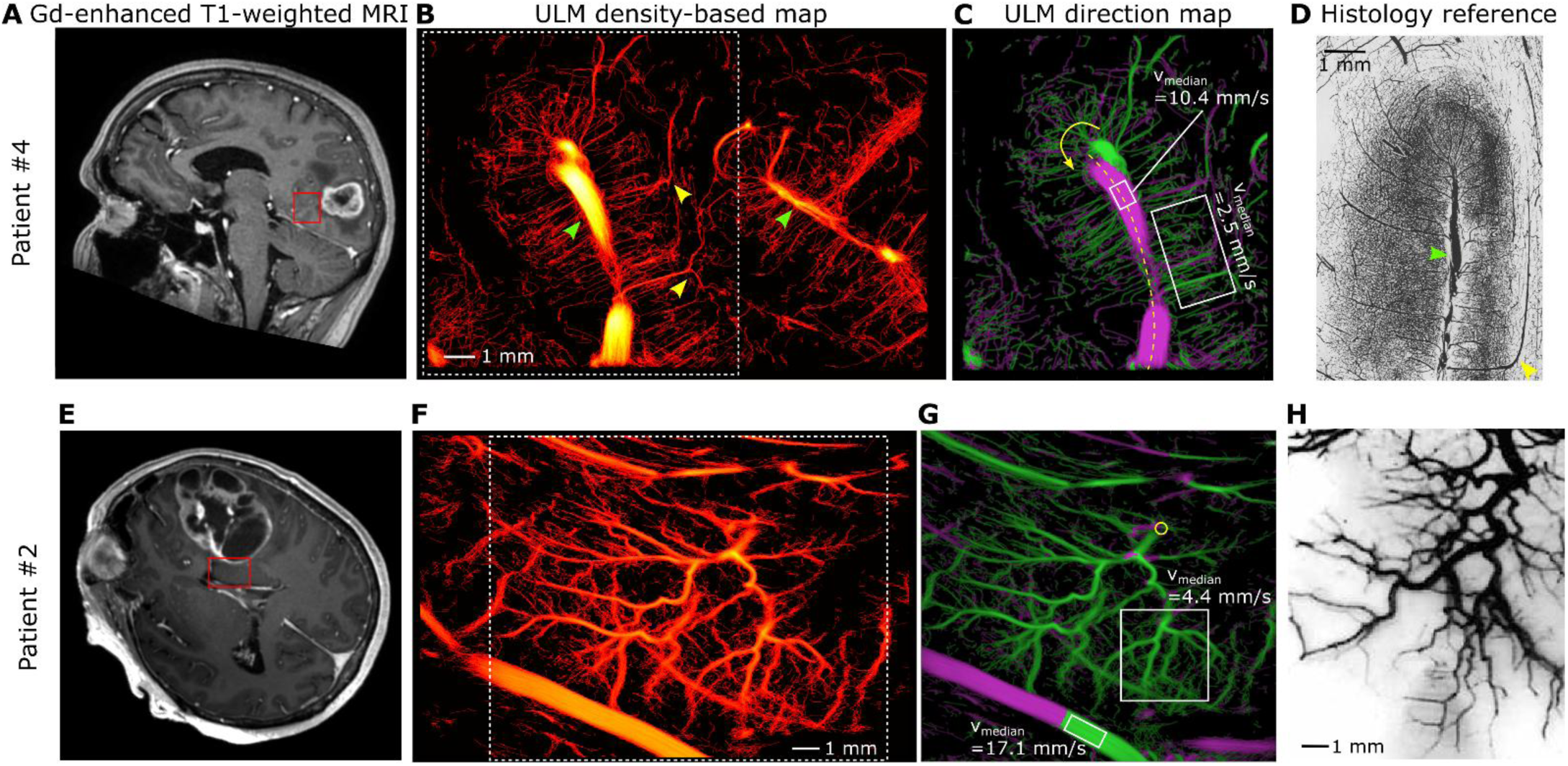
Visualization of microvasculature outside of the tumor. **(A–D)** Cortical vasculature and **(E–H)** striatal vasculature are shown from two example patients: patient #4 (A–C) and patient #2 (E–G). **(A, E)** Contrast-enhanced (Gd) T1-weighted preoperative MR images display the anatomical locations of the ULM regions of interest (ROIs), indicated by red boxes. **(B, F)** ULM intensity maps of the ROIs depicted in (A, E). (B) Shows two intracortical vascular trees consist of pial vessels (green arrowhead) and cortical branches, includng deep cortical branches that extend into the white matter (yellow arrowhead). **(C, G)** ULM direction maps visualize the directionality of microbubble movement away from (green) or towards (purple) a reference line in (C, yellow dashed line) and a reference point in (G, yellow circle). The yellow arrow in (C) indicates the flow direction at the turning point of large pial vessel. **(D)** Ink-stained reference image from Duvernoy *et al* (43) of the vascular network of the central sulcus with the frontal and parietal cortexes visable. An intercortical vessel (green arrowhead) and a deep cortical branch (yellow arrowhead) are labeled. **(H)** Ink-stained reference image from Salamon (47) of the striatal vasculature.

Similar to the first row of Fig. 5, panels E-G show results from patient #2, diagnosed with a metastatic lesion originating from lung carcinoma. In the contrast-enhanced T1-weighted MRI (Fig. 5E), the cystic metastatic tumor can be seen in the left frontal lobe, with the striatum located directly beneath the lesion and highlighted by the red box. ULM was able to capture the vascular pattern within the striatum (Fig. 5F). Median blood flow speeds in two ROIs in Fig. 5G were calculated to be 4.4 mm/s (part of the striatal vasculature), and 17.1 mm/s (an adjacent choroidal vessel). To assess flow directionality, a reference location was selected (yellow circle) at the top of the vascular tree, and microbubble movement away (green) and towards this reference location was calculated (Fig. 5G). Interestingly, blood flow was consistently directed away from the reference, indicating this was a feeding vascular tree. Lastly, the observed branching pattern closely resembled that of canonical striatal vessels, as illustrated in the textbook reference image in Fig. 5H (47). These results show that intraoperative ULM can be used to measure blood flow dynamics at a resolution and depth that was not previously possible, and can contribute to understanding of human brain blood flow on the micrometer scales.

### Variations in microvascular structure across tumor types and patients

Qualitative similarities and differences in microvascular structure were observed between tumor types and patients. Fig. 6 presents a summary of all measured tumors, including both ULM images and their corresponding preoperative MRI slices, on which the ultrasound FOVs are labeled in red. For patient #7, an arbitrary MRI slice is shown, as registration was not possible due to the absence of optical tracking during that measurement. The figure is organized by tumor type: the top row shows three metastases, the middle row displays three high-grade gliomas, and the bottom row shows three meningiomas. Across all recordings, the lateral field of view was 4.5 cm, imaging depth ranged between 6 cm to 8.5 cm, and image resolution was measured to be 42.4 ± 9.2 µm.

**Fig. 6.**
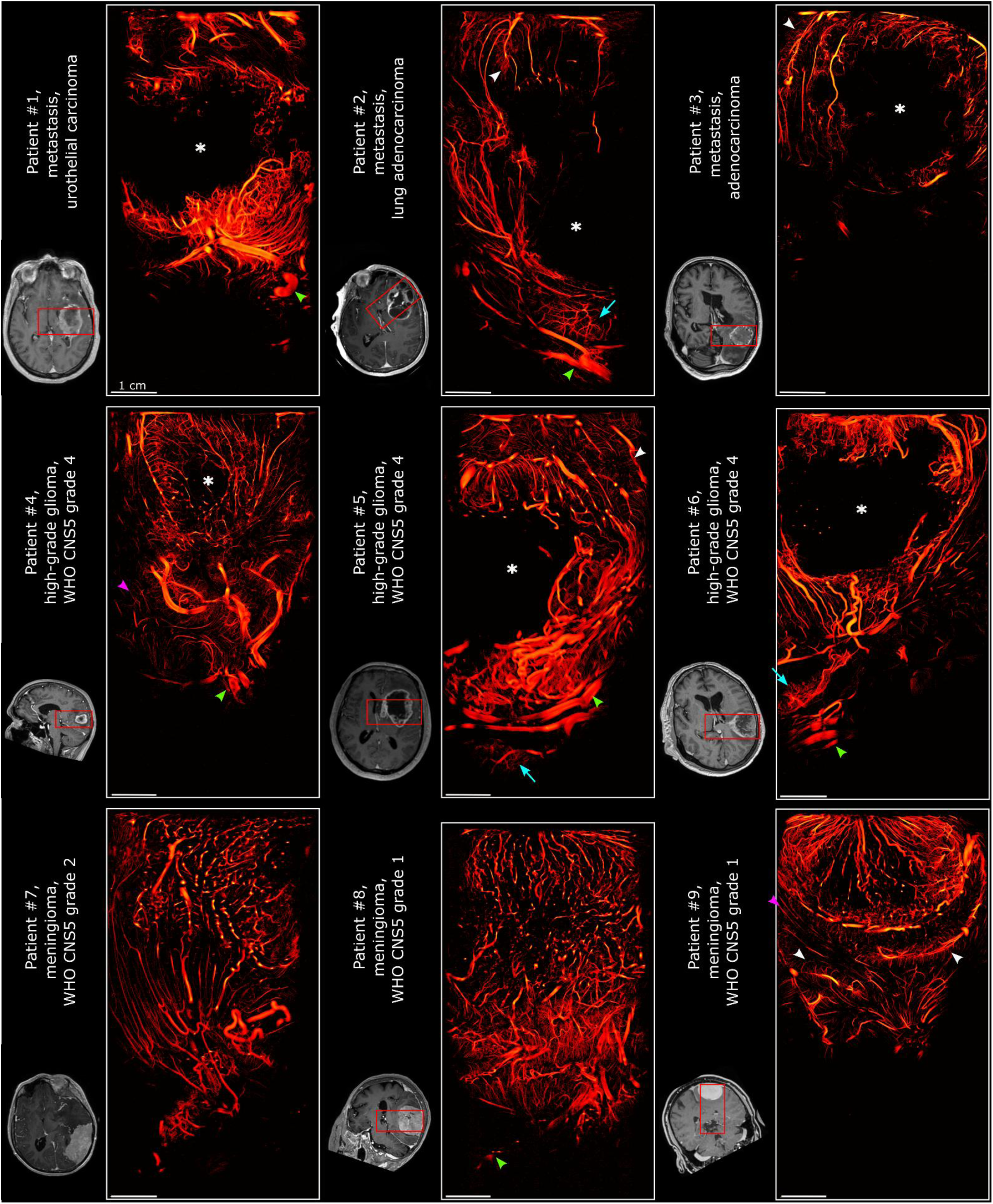
Summary of all 2D measurements. ULM density maps and their corresponding preoperative MRI slices (except for patient #1) are shown. The approximate location of the tumor based on the B-mode image is labeled (white dashed area). The ultrasound FOVs are indicated by red boxes on the MRIs. All images are displayed to 8.5 cm depth. Key landmarks are labeled: necrotic/systic lesions (white asterisk), choroidal vessels (green arrowhead), striatal vessels (turquoise arrows), cortical vessels (white arrowhead), and white matter microvasculature (purple arrowheads).

Different levels of vascularization can be observed across the three tumor types. The meningiomas did not contain any necrotic or cystic regions, whereas the metastases and high-grade gliomas all contained large regions with no blood flow (white asterisks). The vasculatures of the meningiomas in patients #8 and #9 followed the same trend as previously observed with other imaging methods: feeding and draining vessels originate from and drain into vessels at the top of the images, presumably at the meninges. In contrast, no consistent vascular orientation was apparent in the more aggressive tumor types. Structurally, the vessels of the meningiomas were relatively straight, whereas those in the metastases and high-grade gliomas were notably more tortuous—again consistent with previous findings in the literature (8,39).

Interestingly, a major difference existed between patients #8 and #9. Despite both cases being grade 1 meningiomas, patient #9’s tumor exhibited a well-defined border and vasculature that was disconnected from the surrounding brain tissue. In contrast, patient #8’s tumor had a less distinct boundary where the vasculature appeared to be connected. The vascular pattern for patient #8 suggests that this tumor might be more advanced compared to patient #9, despite being categorized with the same grade (grade 1).

Different vascular structures surrounding the tumors could be seen in all recordings. In patients #1,2,4,5,6,8, choroidal vessels were visible (green arrowheads). Deep nuclei microvasculatures were also visible in patients #2, #5, and #6 (turquoise arrows), displaying tree-like structures similar to those previously observed through histology (Fig. 5F). Cortical vessel structures were also found in patients #2, 3, 4, 5, 9 (white arrowheads). Different degrees of modification influenced by the tumor could be seen in these tumors. Vessels resembling the microvasculature of white matter (48) were identified in patients #4 and #9 (purple arrowheads). Directly under the tumor in patient #8, a mixture of compressed gray and white matter led to a densely packed microvascular network that had relatively high blood flow, even compared to the tumor vasculature.

Overall, we observed substantial vascular heterogeneity across tumor types and between patients, although certain common features were present. These findings demonstrate that intraoperative ULM can consistently image microvascular architecture across a range of tumor types and their surrounding tissue.

### AVM structure can be visualized with 3D ULM

While 2D imaging offers valuable insights into microvascular architecture, in the case of AVMs, we trade spatial resolution for 3D visualization, which is essential for capturing their full spatial complexity and for informing precise surgical or endovascular interventions. To this end, we performed three-dimensional ultrafast ULM in patient #10, who was diagnosed with an AVM. Fig. 7 presents the results for this patient. The AVM, with a compact nidus of ∼10.0 x 15.5 x 21.1 mm^3^, was located in the left cerebellar hemisphere of the patient, adjacent to the culmen (Fig. 7A, yellow arrowhead points to the nidus). Preoperative 2D digital subtraction angiography (2D-DSA) identified two primary feeding arteries (Fig. 7B, red arrows) during the arterial phase and a large draining vein (Fig. 7B, right, turquoise arrows) during the venous phase. These feeding vessels were confirmed on preoperative three-dimensional DSA (3D-DSA) (Fig. 7C, red arrows), although a definitive match for the draining vein could not be established with high certainty. Based on its location in 2D-DSA, the most likely draining vessel is annotated in Fig. 7C (turquoise arrow). In both 2D and 3D DSA, vascular structures within the nidus could not be clearly discerned. Ultrafast ULM provided a high-resolution density rendering of the AVM that showed good anatomical overlap with 3D-DSA (Fig. 7D). Although the field of view (FOV) of the ULM acquisition (60° × 60° × 6.0 cm) was smaller than that of 3D-DSA, ULM offered high spatial resolution, estimated at 226 µm using the Fourier shell correlation method (22). Two orientations of the AVM visualized with ULM are shown in Fig. 7E, with draining vessels corresponding to DSA highlighted (white arrows).

**Fig. 7.**
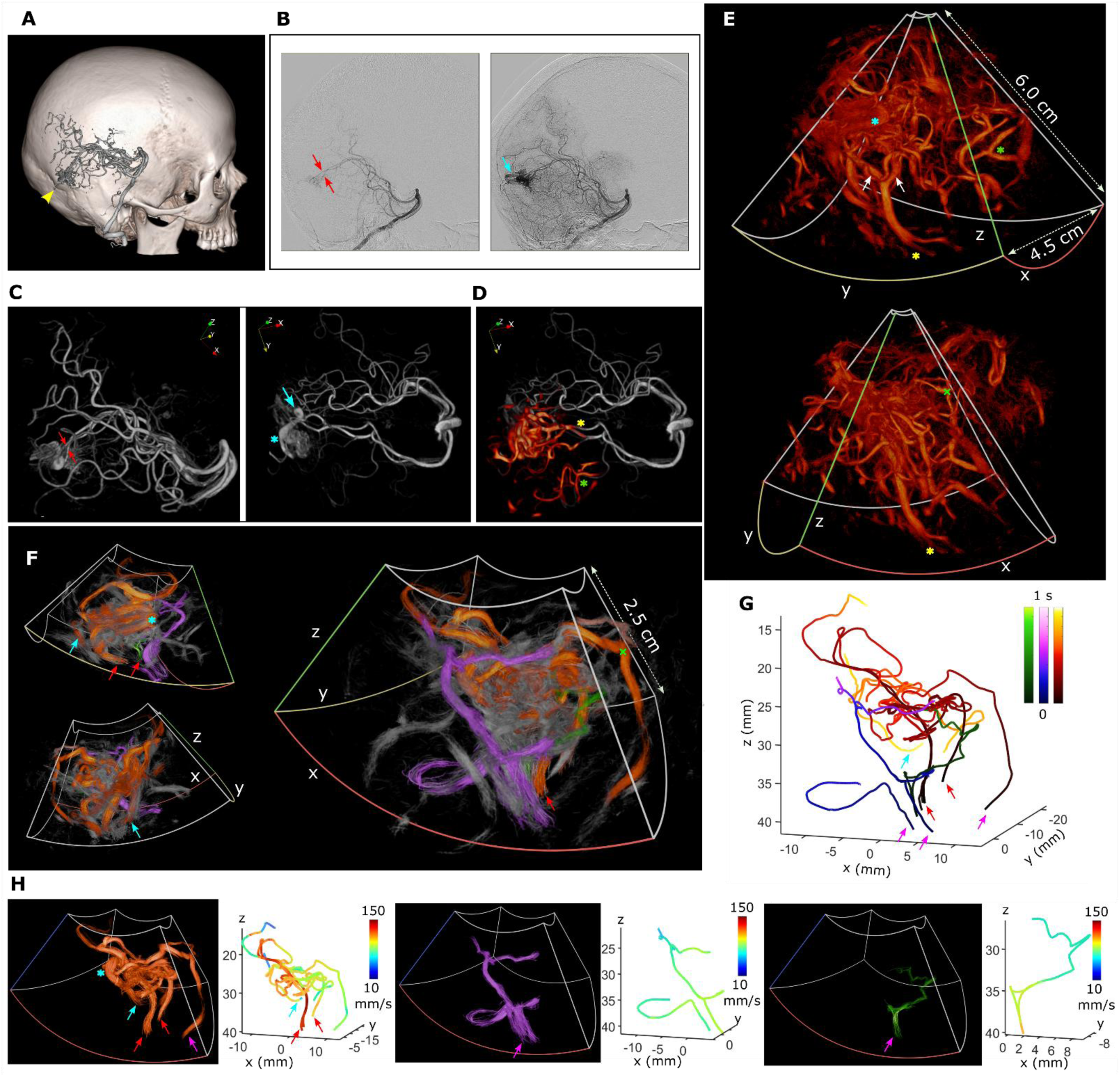
Arteriovenous malformation (AVM) in 3D from patient #10. **(A)** Location of the AVM (yellow arrowhead) relative to the patient skull, demonstrating the cerebellar location of the AVM fed by the posterior circulation. **(B)** Preoperative 2D-DSA images of the arterial (left) and venous (right) phases. Two main feeding vessels (red arrows) and one main draining vessel (turquoise arrow) can be identified. **(C)** The AVM can also be identified in the preoperative 3D-DSA render. The feeding (red arrows) vessels can be identified (left). A small aneurysm can also be seen (right, turquoise asterisk). Due to the limited field of view, the exact same draining vessel as in (B) was not identified. An approximate location, just past the aneurysm, is labeled (right, turquoise arrow). **(D)** Overlay of the ULM (red) and 3D-DSA (gray) images of the AVM. Two vascular structures, a feeding vessel (yellow asterisk) and a peripheral vessel (green asterisk) are labeled to match the same features in (E). **(E)** ULM density-based renders of the AVM in two orientations. A box of size 60° x 60 ° x 6.0 cm is drawn to illustrate the transmit field of the probe. The x, y, and z axes are labeled and color-coded with red, yellow, and green respectively, matching the orientation pointers in (C and D). The aneurysm (turquoise asterisk), a feeding vessel (yellow asterisk, and a peripheral vessel (green asterisk) are labeled as in (C, D). An additional small feeding vessel to the AVM is labeled in (E) and (F) (green cross). (**F)** A zoom-in of the AVM nidus in three orientations. The feeding (red and purple arrows) and draining vessels (turquoise arrow) can be identified. Connected vascular paths are color-coded in orange, purple, and green. Un-tracked vessel paths are in gray. The same vessel features are labeled with asterisks and crosses in (C-E). **(G)** Three main identified vascular units based on vessel connectivity. Color bars present the average flow time starting from the entry points (red and purple arrows). **(H)** Separate renders of the three identified vascular units through the AVM. The blood flow paths are also displayed, color-coded with flow speeds. The feeding vessels (red and purple arrows), a draining vessel (turquoise arrow), and an intranidal aneurysm (turquoise asterisk) are labeled.

Zooming in on the AVM nidus, 3D visualizations of the ULM speed-based volumes reveal complex vascular structures within the lesion (Fig. 7F). By tracking individual microbubbles (see Methods: 3D blood vessel segmentation), we identified three major vascular pathways that supplied and drained the nidus (Fig. 7F, color-coded vessels). Using microbubble position, directionality and speed, we reconstructed these vascular components and computed traversal times through the nidus (Fig. 7G). Separate 3D visualizations of each path and their corresponding flow trajectories are shown in Fig. 7H. The most complex path (orange) involved multiple feeding arteries branching from the main arterial feeder (red and purple arrows), including the two feeding vessels previously observed with DSA (red arrows). Moreover, a small intra-nidal aneurysm (turquoise star) and a primary draining vein (turquoise arrow) could also be identified. While a main draining vessel was detected, its overlap with DSA imaging could not be confirmed with 100% accuracy due to the limited ULM field of view. For the orange pathway, microbubbles traversed its length in up to 0.9 seconds. For the purple and green pathways, feeding vessels were clearly delineated (purple arrows), but no definitive draining veins were observed, as the tracked vessels tapered in diameter and bubble count until the path could no longer be reliably followed. Flow speeds along each identified pathway are color-coded and displayed in 3D (Fig. 7H, right). Across all pathways, measured velocities ranged from 26.3 mm/s to 147.9 mm/s. These results demonstrate that intraoperative 3D ULM can resolve the intricate architecture and dynamic flow patterns of AVMs with high spatial and temporal precision, offering a potential tool for detailed surgical planning beyond the capabilities of conventional angiography.

## DISCUSSION

Intraoperative ULM improves upon traditional intraoperative Doppler and other imaging modalities by offering a range of hemodynamics-based biomarkers: sub-millimeter vascular resolution, quantification of blood flow dynamics, assessment of vascular connectivity, and the ability to resolve feeding, draining, and internal structures of the AVM nidus. In this study, we demonstrated the feasibility of intraoperative ULM in 10 patients with various neurosurgical pathologies, achieving strong correspondence between ULM-derived vascular maps and preoperative MRI or 3D DSA. Notably, ULM enabled the visualization of microvascular features that were undetectable on MRI and, in some cases, previously only observable through histology. (5,6,8). The additional blood flow information, previously unavailable at such high spatial and temporal resolution, can potentially open new avenues for the discovery of vascular biomarkers and may improve tumor characterization.

In addition to characterizing tumor vasculature and hemodynamics, intraoperative ULM offers valuable insights into vessel connectivity, which may directly inform surgical decision-making during tumor resection. By tracking individual microbubbles as they traverse the vasculature, ULM enables the visualization of vascular pathways with high fidelity, allowing differentiation between vessels that feed or drain the tumor and those merely adjacent to or bordering it. This level of detail—combined with blood flow information—can help guide the surgeon in identifying optimal resection strategies to minimize blood loss and preserve surrounding healthy tissue (13,14). In the AVM, we did not reach the same high resolution (226 µm in 3D vs 35 µm in 2D) due to the lower center frequency, lower PDI resolution in 3D, probe motion, and shorter recording durations. Nevertheless, we achieved three-dimensional visualization of the compact nidus structure, along with its feeding and draining vessels—critical information for planning and executing microsurgical interventions (49). Moreover, ULM’s ability to capture flow dynamics within the nidus offers an added layer of functional detail, potentially refining surgical strategies. In both the tumor and the AVM, intraoperative ULM delivered spatial and dynamic vascular information that exceeded the capabilities of conventional intraoperative imaging modalities.

Despite its promising capabilities, intraoperative ULM faces several limitations that currently hinder its routine adoption in surgical practice. One major constraint is its inherently limited field of view compared to modalities such as MRI or X-ray angiography, which provide broader anatomical context. As a result, ULM often relies on these modalities for orientation and spatial reference. This limitation can be mitigated by employing ultrasound probes with larger opening angles or by performing consistent live registration with preoperative imaging—an approach we already incorporate intraoperatively for Doppler (30). A current obstacle for wide-spread clinical use is that the post-processing workflow is complex, time-intensive, and requires expert oversight. Standard ULM image reconstruction typically takes hours for 2D up to a day for 3D, with further delays introduced by the need to fine-tune parameters for individual patients. Many aspects of the analysis remain semi-automatic, making trained interpretation essential and limiting scalability. However, ULM is a rapidly advancing field, with ongoing developments in algorithm acceleration, GPU-based parallelization, and deep learning-based processing (50,51). These innovations are steadily pushing ULM toward real-time or near-real-time implementation, significantly enhancing its potential for clinical integration and broader use in neurosurgical procedures.

Another major challenge (and perhaps the most important) for broader clinical application of ULM is the skull. The skull significantly reflects, attenuates, and aberrates ultrasound waves, making it difficult to achieve the high spatial resolution and image quality demonstrated in this study without craniotomy (52,53). Fortunately, research is actively ongoing to improve aberration correction techniques, which—when combined with appropriately designed transducers and strategically selected acoustic windows—could make transcranial ULM feasible. Recent studies have already reported promising results in this direction (31,54,55). Another avenue for bypassing the skull would be the combination of ULM with imaging through cranioplasty, replacing human skull bone, as recently demonstrated for non-contrast enhanced ultrafast ultrasound (56,57). Successful adaptation of transcranial ULM for clinical use could profoundly reshape the diagnostic workflow for brain lesions. Given that contrast-enhanced ultrasound is radiation free, significantly more cost-effective, and more accessible than MRI, CT, or X-ray angiography, this shift could enable broader and earlier detection of neurovascular abnormalities, particularly in resource-limited settings.

In this study, we included a variety of tumor types to demonstrate the feasibility of intraoperative ULM in visualizing tumor vascular structures. As a next step, larger-scale studies focusing on individual tumor types are warranted to enable meaningful quantitative analyses and comparisons across tumor subtypes. An additional area of interest is the assessment of blood flow in deep brain regions, where established imaging modalities often lack the sensitivity to detect slow microvascular flow at high spatial resolution. Improved integration of optical tracking systems with 3D ultrasound could also enhance spatial accuracy and image reconstruction, potentially reducing or eliminating the need for complex motion compensation techniques. Ultimately, intraoperative ULM holds great promise not only for advancing our understanding of human brain lesion vasculature, but also for contributing directly to surgical decision-making by providing real-time, high-resolution vascular information.

## MATERIALS AND METHODS

### Study design and patient recruitment

The objective of this study was to evaluate the feasibility of intraoperative ULM for imaging human brain tumor microvascular structure and dynamics, characterize vascular features across different tumor types, and assess its potential to inform surgical decision-making.

Ethical Approval Prior to inclusion, written informed consent was obtained from all patients in line with the Dutch national medical-ethical regulations as formulated by The Central Committee on Research Involving Human Subjects (CCMO) in the Netherlands. The Erasmus Medical Center’s Medical Research Ethics Committees (MREC), located in Rotterdam, the Netherlands, reviewed the three research protocols underlying the current study and ethical approval was given on: intraoperative functional Ultrasound (fUS)-imaging during awake and anesthetized neurosurgical procedures (fUS Studie), protocol number MEC-2018-037 (Ethical approval was given on 22nd of January 2020). All methods in this study were carried out according to the protocol. All subjects gave permission for the use and publication of their images and data. Participants were eligible for inclusion if they were over 18 years of age, had been diagnosed with a brain tumor and AVM scheduled for surgical removal under general anesthesia, and had no contraindications to Sonovue (Bracco, Italy).

### Study procedure

Ultrasound recordings were performed after craniotomy. The ultrasound probe (both 2D and 3D) was placed in a sterile sleeve, pre-filled with ultrasound gel, and was placed on top of the intact dura above the tumor. Saline was added to the operating field to ensure adequate acoustic coupling. The 2D imaging probe was fixed in place by a custom-made articulated arm for patients #1-9. Probe fixation was not possible for patient #10 (3D imaging) due to the small size of the craniotomy and the depth to the dura (thick muscle layers at the back of the head).

Sonovue was prepared according to the package instructions by the anesthesiologist approximately 10 minutes prior to injection. Each bolus with 0.1 to 0.4 mL volume was slowly injected by hand into a venous line with a syringe. Directly after contrast injection, a saline flush of 10 mL was injected into the venous line with a programmed duration of 30 s. The total volume of contrast used for each measurement was under 1 vial (5 mL).

Synchronized with the contrast injections, 3.5-minute or 3-minute duration ultrasound recordings were acquired for 2D and 3D respectively, capturing the contrast bolus arrival and part of the circulation time. In total, between 1 and 4 recordings were made to build one ULM image. The ultrasound probe was tracked using the neuro-navigation system (Curve, BrainLab, Germany) to allow registration of ultrasound and preoperative MRI/CT images during post-processing. Tracking was not performed in patients #7 and #10 due to the lack of tracking hardware and limited space around the probe to attach the tracking star respectively. The entire measurement extended the conventional surgical procedure by a maximum of 22 minutes. The collected beamformed data was stored and processed offline.

### 2D imaging parameters

2D ultrafast ultrasound recordings were performed using a programmable research ultrasound system (Vantage 256, Verasonics, USA) interfaced with a linear array (GE9L-D, GE, USA). The transmission sequence consisted of 2-cycle pulses centered at 5.2 MHz. Angled plane waves (10 angles equally spaced between −6° and 6°) were transmitted at a pulse-repetition frequency of 6 kHz (duty cycle = 0.23%). The acoustic amplitude and intensities (MI = 0.1 and ISPTA = 44.3 mW/cm^2^) were largely below the FDA recommendations (MI = 1.9 and ISPTA = 720 mW/cm^2^). The received signals were sampled at 100% bandwidth, beamformed using the Fourier beamformer to a pixel size of 0.5 lambda, and saved to an SSD drive. The entire process was performed in real time with custom software. Our system has the capability to save continuous IQ data for an arbitrary amount of time (or until no more disc space was available), and in this case we performed acquisitions of 3.5 minutes in duration.

### 3D imaging parameters

3D ultrafast ultrasound recordings were performed using the same programmable research ultrasound system (Vantage 256, Verasonics, USA) interfaced with a trans-esophageal (TEE) matrix array (Oldelft Ultrasound, the Netherlands). A detailed description of the probe can be found in (58). The probe has an aperture of 11.52 x 8.64 mm, consisting of 3072 (64×48) elements organized into groups of 4×4 for analogue micro-beamforming, thus reducing the channel count to 192. The transmission sequence consisted of 3-cycle pulses centered at 3.6 MHz, with 16 partially overlapping diverging beams making up a FOV of 60 ° x 60°. The pulse repetition frequency (PRF) was 4 kHz, leading to a frame rate of 250 Hz. The acoustic amplitude and intensities (MI < 0.4 and ISPTA < 88.2 mW/cm^2^) were also below the FDA recommendations. The received RF data was sampled at 100% bandwidth and beamformed using our custom GPU-based DAS beamformer into the spherical domain. The resulting IQ data were used for further ULM post-processing. In total, three recordings at different locations were performed, of which two were used (one did not have a good view of the AVM).

### 2D ULM post-processing

The beamformed IQ data were filtered for the microbubbles, which were localized and tracked to yield ULM maps of the tumor and surrounding tissue. For the 2D images, clutter filtering was performed using the SVD filter (59) for ensembles of 1000 frames. The first ranks (between 10 and 30) that consist mostly of tissue signals were determined manually for every recording and were removed. Microbubble localization, tracking, and ULM map visualization were performed using the PALA toolbox (34), with a few modifications including localization by Gaussian fitting, noise thresholding, TGC gain, lag-1 auto-correlation for noise rejection, and Makima interpolation for continuous tracks (60). Identified tracks from different recordings of the same patient were merged before rendering. In all recordings, interframe motion was minimal, so no motion correction was performed. By tracking single microbubbles, their positions in time were obtained and used to calculate for movement direction, speed, density per pixel etc., rendered at 0.015 mm pixel size. Fig. S2 contains a flow chart that outlines these processing steps. For Fig. 3, the distance metric for track segments was calculated as

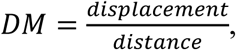

while keeping the segment length or distance to 1 mm, similar to (40).

Relative blood volume of any region was calculated by

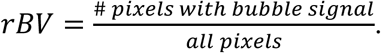

### 3D ULM post-processing

The received RF data were beamformed using a custom delay-and-sum algorithm (61) onto a spherical grid spanning 90° in both azimuth and elevation (wider than the transmission angle of 60°), with a depth range of 6.0 cm. The grid had an angular resolution of 0.94° and a depth sampling interval of 0.5 mm. The beamformed data were split into ensembles of 384 frames each (212 ensembles in total) and processed using the PALA toolbox, modified for 3D data to include processes in (60). The output from the ULM processing included all microbubble trajectories (location and velocity) rendered at 0.086 mm^3^ voxel size in the cartesian domain.

Because the ultrasound probe was hand-held by the surgeon during the minute-long recordings, the data contained significant inter-ensemble motion. To estimate motion, ULM maps were reconstructed for each ensemble, and rigid affine transformations (62) Cumulative motion was estimated by aligning consecutive frames, followed by a second-pass global registration that used the midpoint of the sequence as a reference to correct residual misalignments. The resulting motion parameters were used to correct microbubble trajectories and reconstruct a final 3D ULM volume. For multi-volume registration, a rigid transformation was computed (imregtform, Image Processing Toolbox, MATLAB R2024a) followed by a non-rigid transformation (imregdemons, Image Processing Toolbox, MATLAB R2024a) to align the two volumes. Fig. S2 contains a flow chart that outlines these processing steps. Finally, two volumes were registered for the ULM images shown in Fig. 7.

### Probe tracking and multi-modality registration

For 2D measurements, optical tracking of the ultrasound transducer, combined with standard patient tracking, enabled the alignment of ULM data with preoperative imaging modalities. Details of the setup and processing are described in references (30,63). To further refine the registration, translational manual adjustments (within 5 mm) were performed in 3D Slicer (64) to improve matching between major landmarks and the overall registration.

For 3D measurements, optical tracking was not feasible due to limited space in the craniotomy, which prevented attachment of the probe tracking marker. Instead, all registrations were performed offline. To align the ULM volume with the preoperative 3D-DSA (Fig. 7), the rasterized ULM volume was first down sampled by a factor of four in all directions to match the resolution of the 3D-DSA. An initial manual alignment was then performed in ParaView (Kitware, USA) by adjusting translation and rotation. Next, the two volumes were registered by estimating their rigid displacement (imregtform, Image Processing Toolbox, MATLAB R2024a). A second round of non-rigid motion correction was also performed (imregdemonsm Image Processing Toolbox, MATLAB R2024a), accounting for any deformations of the brain that occurred post-craniotomy.

### 2D blood vessel segmentation

Segmentation of vessels was performed for Fig. 4C-E using a semi-manual custom script developed in MatLab. First, ULM processing was performed to generate all microbubble tracks. To begin segmentation, a vessel of interest was selected by identifying an initial query point in a region without overlapping or crossing vessels. Two points were then manually defined on either side of the vessel near this query point to delineate its approximate boundaries. All microbubble tracks passing between these two boundary points were selected as part of the vessel. Based on the spatial continuity and connectivity of these tracks, the next query point was chosen along the same vessel or a branching segment. This stepwise process was repeated iteratively until the entire vascular tree of interest was reconstructed.

### 3D blood vessel segmentation

Segmentation of the blood vessels in the AVM was performed using custom semi-automatic software developed in MATLAB. The software is designed to iteratively follow microbubble tracks in incremental spatial steps, classifying track clusters that belong to the same vessel and continuing along the vessel path until track density becomes insufficient for further progression.

First, ULM processing was performed to generate all microbubble tracks. The process began by manually selecting a starting location, typically a large, easily identifiable vessel near the edge of the field of view, based on visual inspection of the 3D dataset. A representative starting speed was also defined at this location. The algorithm then searched for all bubble tracks within 4 mm of the starting point that had a direction vector within 80° of the initial direction. These tracks were averaged to create a reference trajectory. From this reference, the algorithm advanced 3 mm along the vessel path and identified nearby tracks satisfying the same spatial and directional constraints. For each step, it computed and stored the position and velocity of candidate tracks at both the current and next locations, forming a 9-dimensional feature space (e.g., position, direction, and velocity). The optimal number of k-mean clusters within this space was then determined by maximizing the silhouette value with varying number of clusters (1 to 5). All resulting clusters were displayed for manual review, and the clusters corresponding to the vessel of interest were selected. A new reference track was computed based on these selected tracks, and the algorithm advanced another 3 mm along this new path. This iterative process continued until the algorithm reached a point where the remaining tracks were too sparse or too short to reliably define the vessel path.

### Statistical analysis

Wilcoxon rank-sum tests were performed in MATLAB 2022b to evaluate for statistical significance of DM and speed of the tumor and non-tumor regions (Fig. 3). Statistical significant differences were observed (P<0.001).

## List of Supplementary Materials

Table S1. Overview of patient information.

Fig. S1. ULM density maps overlaid on preoperative MRIs of three example tumors. Fig. S2. Flow charts of 2D and 3D ULM processing steps.

Video S1. Microbubble tracking for the caudate microvasculature (patient #2).

Video S2. Overview of all 2D ULM datasets.

Video S3. Rotational render of the AVM (patient #10).

## Data Availability

All data produced in the present work are contained in the manuscript.

## Acknowledgments

We would like to thank the neurosurgical anesthesiology team for neurosurgery, the operating room team, and the operating room technical team of the Erasmus MC in Rotterdam, including Jolanda van den Berg, Josiane Wink-Godschalk, Jan van Beest, and Michael Smits. Additionally, we would like to thank Fits Mastik, Stefanos Florescu, Jason Voorneveld, and Yichuang Han for their technical discussions and support. During the preparation of this manuscript, the authors used *ChatGPT-5* (OpenAI) and *Claude* (Anthropic) to assist with language polishing and improving readability. The authors reviewed and edited all content generated by this tool and assume full responsibility for the final text.

## Author contributions

Conceptualization: LW, PK Methodology: LW, LV, PX, ECG, PPS

Investigation: LW, LV, ECG, SS, WBLB, VV, CMFD, JWS, EMB, AK, PPS, AJPEV

Visualization: LW, PK Funding acquisition: PK, MS Project administration: LW, PK

Supervision: AJPEV, PK Writing – original draft: LW, PK

Writing – review & editing: LW, LV, PX, SS, MS, VV, EMB, PPS, AJPEV, PK

## Data and materials availability

All data and code associated with this study will be made available.

