## Supplementary materials for "Intraoperative ultrasound localization microscopy of human brain tumors and arteriovenous malformations"

Table S1. Overview of patient information.

| | Age range (Sex) | Tumor type | Location |
|---|---|---|---|
| **Patient #1** | 66-70 (M) | Metastasis, urothelial cell carcinoma | Left parietal |
| **#2** | 66-70 (F) | Metastasis, lung adenocarcinoma | Left frontal |
| **#3** | 71-75 (M) | Metastasis, colorectal adenocarcinoma | Left parietal |
| **#4** | 61-65 (F) | High-grade glioma, WHO CNS5 grade 4 | Left occipital |
| **#5** | 61-65 (M) | High-grade glioma, WHO CNS5 grade 4 | Left frontal |
| **#6** | 66-80 (M) | High-grade glioma, WHO CNS5 grade 4 | Left parietal |
| **#7** | 46-50 (F) | Meningioma, WHO CNS5 grade 2 | Left parietal |
| **#8** | 76-80 (F) | Meningioma, WHO CNS5 grade 1 | Left parietal-occipital |
| **#9** | 46-50 (M) | Meningioma, WHO CNS5 grade 1 | Left frontal-parietal |
| **#10** | 61-65 (F) | AVM S-M gr II cerebellar left inferior confluens sinuum | Left cerebellum |

**Intraoperative ULM reveals unresolved microvascular structures in MRIs**

To spatially align the ultrasound images with the other imaging modalities, the position and orientation of the ultrasound probe were tracked using an optical tracking system (BrainLab, Germany) during the operation (15). The tracking data was used to find the corresponding 2D slices on the preoperative MRIs (see Methods: probe tracking and multi-modality registration).

Figure S2 shows three examples of ULM density-based maps alongside their corresponding MRI slices for three tumor types: a metastasis (Fig. S2A and B), a high-grade glioma (Fig. S2C and D), and a meningioma (Fig. 2E and F). The ultrasound field of view (FOV) was 4.5 cm x 8.5 cm (Fig. S2A-F, left). In these examples, vessels were visualized down to a depth of 6 cm.

Good spatial overlap was achieved between ULM and MRI in all three cases. Larger vessels, such as the choroidal vessels in panels A–C (yellow arrows), a pial vessel in panel A (purple arrowheads), and tumor-surrounding vessels in panels A and F (green arrowheads), were clearly resolved in both ULM and MRI images. The close correspondence of these features confirms successful registration between the two modalities.

Tumor centers that appeared hypointense on contrast-enhanced T1-weighted MRI also showed an absence of vessels on ULM (yellow asterisks), consistent with the presence of necrotic cores (Fig. S2A–D). Conversely, hyperintense regions on T1-weighted gadolinium-enhanced MR images corresponded well with highly vascularized areas on ULM, suggesting these vessels are likely the highly permeable vasculature typical of aggressive tumors. In the T2 hyper-intense regions outside of the tumors, nearly no microvasculature was found in the metastasis case (Fig. S2B) whereas some vasculature was detected in the high-grade glioma (Fig. 2D). Additionally, in both the high-grade glioma and meningioma cases, ULM revealed vascular structures resembling healthy cortical vessels (turquoise arrows in Fig. S1C and E). These vascular features also corresponded spatially with gray matter on the MRI scans.

Overall, the strong concordance between ULM maps and MRI images validates the accuracy of ULM in depicting vascular structures. Moreover, the superior spatial resolution of ULM allowed visualization of microvascular details that were not detectable on MRI.

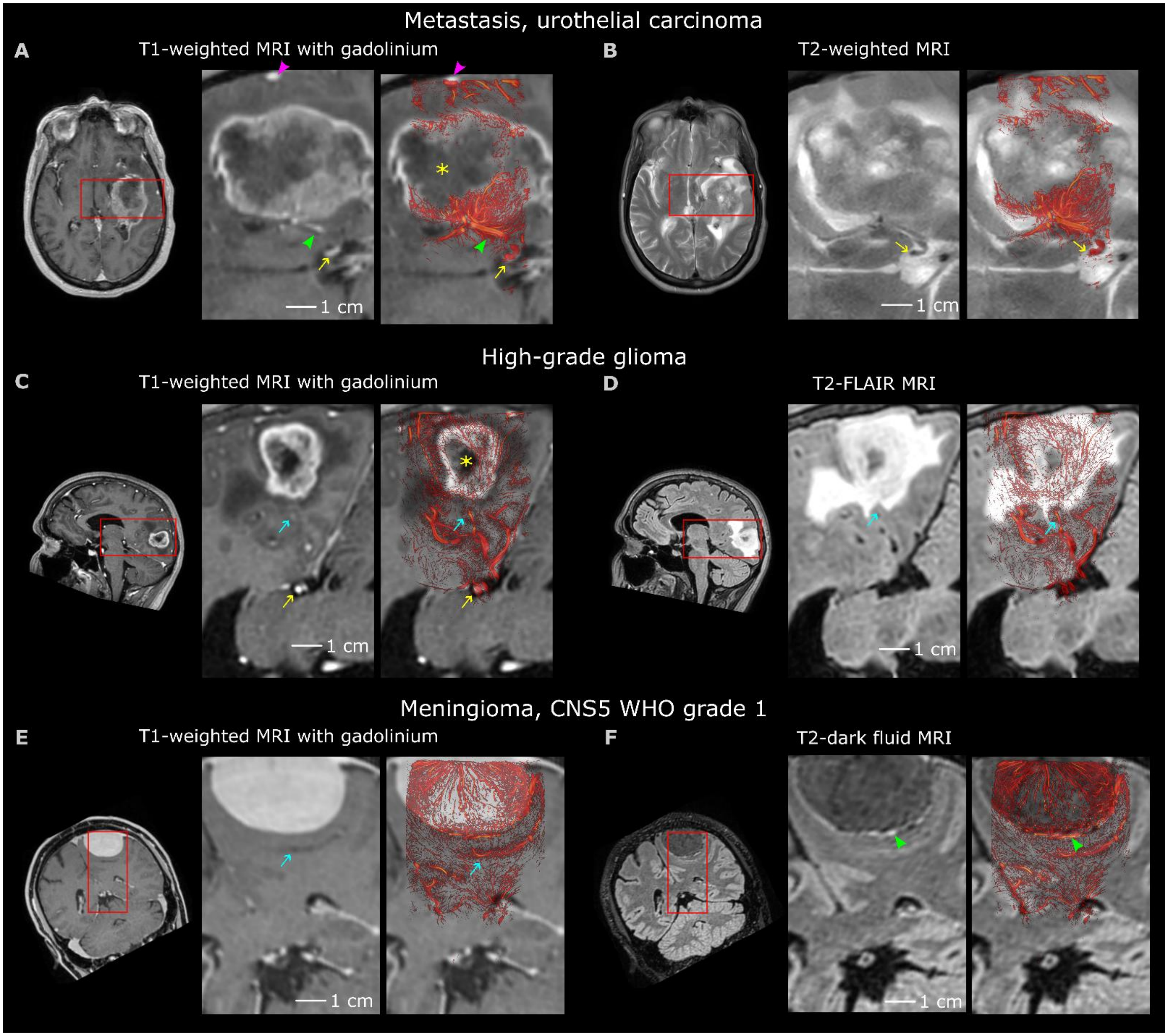


**Fig. S1. ULM density maps overlaid on preoperative MRIs of three example tumors. (A, B)** An example of a metastasis (patient 1). **(C, D)** An example of a high-grade glioma (patient 54. **(E, F)** An example of a meningioma (patient 9). The ultrasound field of view is labled in red on the MRIs (left). Both T1-weighted (A, C, E) and T2-weighted (B, D, F) MR images are included. Certain features are labeled including the tumor nechrotic core (yellow asterisk), choroidal vessels (yellow arrows), superficial pial vessels (purple arrowheads), tumor-surrounding vessels (green arrowheads), and cortical microvasculature (turquoise arrows).

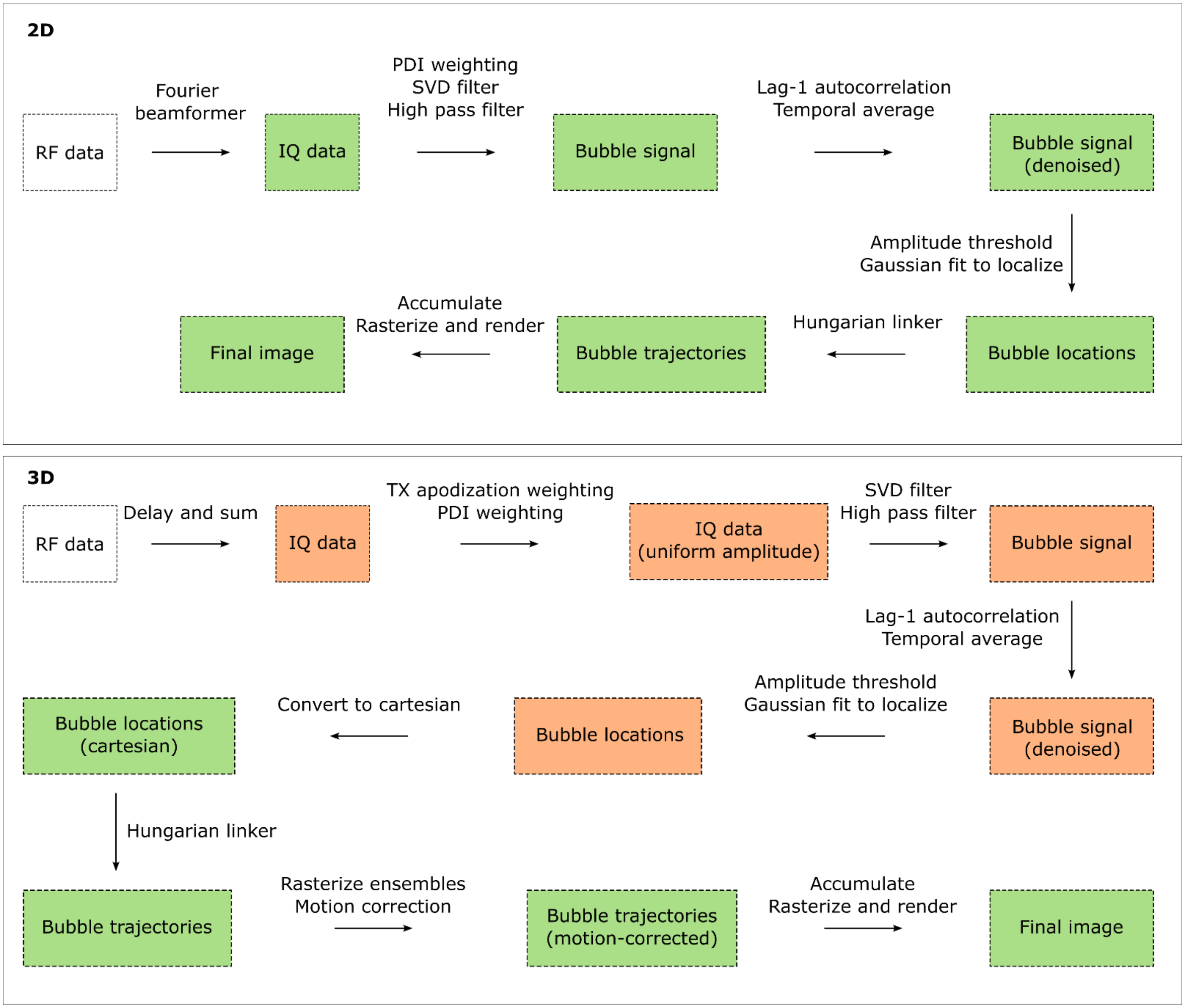


**Fig. S2. Flow charts of 2D and 3D ULM processing steps.** Abbreviations include: RF: radio-frequency; IQ: in phase and quadrature; SVD: singular value decomposition; TX: transmit; PDI: power Doppler image. Steps performed in spherical and cartesian domains are labeled orange and green respectively.

### Supplementary videos

#### Video s1. Microbubble tracking for the caudate microvasculature (patient #2)

Focusing on the caudate nucleus located below the tumor in patient #2, this video shows localized microbubble locations (yellow) overlaid on top of B-mode frames. Over the 20s of accumulation time in this example, the feeding microvascular structure of the caudate nucleus is visualized.

#### Video s2. All 2D ULM density, flow direction, and MRI overlay results.

ULM density and z-direction maps as well as their corresponding B-mode and pre-operative MRI/CT images are displayed here for all tumor patients.

#### Video s3. Rotational render of the AVM (patient #10)

Rotational render of the AVM corresponding to Fig. 7 G, H as well as the three identified vascular paths. The x, y, and z axes are labeled in colors red, yellow, and green respectively. For scale, the length of the green z axis is 2.5 cm.
